# Early Prognostication in Non-Reperfused Stroke: Integrating FVH with CT Perfusion for Personalized Management

**DOI:** 10.1101/2025.11.13.25340185

**Authors:** Qiushi Yang, Yanxin Ma, Xin Qu, Yu Chen, Hui Sun, Gang Yao

## Abstract

**Background/Objectives:** Acute ischemic stroke (AIS) is the second leading cause of death globally, with outcomes heavily dependent on collateral circulation. This study investigated the prognostic value of integrating fluid-attenuated inversion recovery vascular hyperintensity (FVH) from MRI with CT perfusion (CTP) parameters for personalized management of non-reperfused AIS patients, focusing on the correlation between FVH and CTA-based collateral scores (rLMC, Maas, ASITN/SIR).

**Methods:** We retrospectively enrolled AIS patients with internal carotid artery/middle cerebral artery stenosis/occlusion who did not receive reperfusion therapy within 72 hours of onset. All patients underwent one-stop CTA-CTP and multimodal MRI to evaluate: FVH scores (based on modified ASPECTS regions), rLMC scores, Maas scores, and ASITN/SIR collateral grading. Spearman analysis assessed correlations between FVH and CTA collateral scores. Univariate and multivariate logistic regression identified independent predictors of 90-day functional outcome (favorable [mRS 0-2] vs. poor [mRS 3-6]), with Receiver operating characteristic (ROC) curves evaluating predictive performance.

**Results:** The cohort comprised 112 patients (70 favorable outcomes, 42 poor outcomes). FVH scores showed negative correlation with ASITN/SIR collateral grades (r=-0.432, P<0.001). Compared to the favorable outcome group, the poor outcome group exhibited higher baseline National Institute of Health Stroke Scale (NIHSS) scores, elevated FVH scores, reduced rLMC scores, and lower rCBV values (all P<0.05). Multivariate analysis identified NIHSS score, FVH score, rLMC score, and rCBV as independent predictors of poor outcomes. ROC analysis demonstrated strong predictive performance for: rLMC score (AUC=0.848, 95%CI 0.778-0.919), FVH score (AUC=0.662, 95%CI 0.550-0.774), and rCBV (AUC=0.727, 95%CI 0.631-0.822).

**Conclusions:** Multimodal CT combined with MRI facilitates early AIS diagnosis and collateral assessment. The integration of FVH with CT parameters (rLMC score, rCBV) effectively predicts functional outcomes in AIS patients.

## 1. Introduction

A Global Burden of Disease (GBD) study revealed a substantial annual increase in stroke incidence and mortality, establishing stroke as the world’s second-leading fatal disease after cardiovascular disease, characterized by its high morbidity, disability rate, and mortality. Among all stroke subtypes, acute ischemic stroke (AIS) caused by large- or medium-artery stenosis or occlusion is the most prevalent [1].

Clinical studies have observed significant variability in neurological recovery and long-term outcomes among AIS patients with similar degrees of intracranial large-artery stenosis or occlusion, even after receiving thrombolysis or mechanical thrombectomy [2-5]. Emerging evidence suggests that collateral circulation formation and compensatory capacity are critical determinants of ischemic penumbra survival and clinical prognosis [6]. When blood flow is interrupted due to arterial occlusion or stenosis, pre-existing anastomotic vessels dilate or form capillary connections under hypoperfusion stress, establishing microcirculatory bypasses that redirect blood flow to ischemic regions [7] .

Conventionally, collateral status is assessed using computed tomography angiography (CTA)-based scoring systems. CTA offers a noninvasive, cost-effective, and reproducible method for collateral evaluation, providing multimodal information crucial for AIS diagnosis, treatment selection, and prognosis [8] .

In 1999, Cosnard et al. [9] first described fluid-attenuated inversion recovery vascular hyperintensity (FVH) on MRI-FLAIR sequences—a serpentine hyperintensity along cortical vessels, typically in the frontal or temporal lobes. FVH likely reflects slow anterograde or retrograde collateral flow distal to stenotic/occluded vessels due to diminished flow-void effects, serving as a noncontrast marker of leptomeningeal collaterals in early AIS. Studies present conflicting findings: Lyu et al. [10] identified FVH as an independent predictor of poor functional recovery, suggesting its association with inadequate collaterals, whereas Xin et al. [11] correlated higher FVH scores with robust collateral grades and favorable outcomes.

Although multimodal MRI remains sensitive for early infarct detection, prolonged scan times and motion artifacts limit its utility. Conversely, one-stop CTA-CTP provides rapid, comprehensive hemodynamic assessment but may miss early ischemic changes. Combining these modalities could improve prognostic accuracy by evaluating collateral status and perfusion dynamics [12].

## 2. Materials and Methods

### 2.1 Study Design and Participants

We conducted a retrospective analysis of 224 AIS patients admitted to the Department of Neurology, Daqing Oilfield General Hospital, from May 2023 to December 2024. After strict screening per inclusion/exclusion criteria, 112 patients were enrolled. The study protocol was approved by our Institutional Review Board, and written informed consent was obtained from all participants in accordance with the Declaration of Helsinki.

### 2.2 Clinical Data Collection

We collected comprehensive baseline clinical data from all enrolled patients, including gender, age, admission-to-examination time interval, systolic and diastolic blood pressure, heart rate, stroke risk factors (hypertension, diabetes mellitus, hyperlipidemia, coronary artery disease, atrial fibrillation, previous stroke history), location of arterial stenosis or occlusion, and serological biomarkers (total cholesterol [TC], triglycerides [TG], high-density lipoprotein [HDL], low-density lipoprotein [LDL], homocysteine [HCY], uric acid [UA], blood glucose [BS], and glycated hemoglobin [HbA1c]). Neurologists assessed National Institutes of Health Stroke Scale (NIHSS) scores on admission day. The modified Rankin Scale (mRS) was used to evaluate 90-day clinical outcomes, with patients categorized into favorable outcome (mRS 0-2) and poor outcome (mRS 3-6) groups.

#### 2.2.1 Inclusion Criteria

1. Age ≥18 years.
2. Time from symptom onset to completion of one-stop CTA-CTP and multimodal MRI ≤72 hours.
3. Diagnosis of AIS confirmed by DWI or CTA, with severe stenosis (≥70%) or occlusion of the internal carotid artery (ICA) and/or middle cerebral artery (MCA).
4. Received standard medical therapy (antiplatelet/anticoagulant, statins, risk factor control).

#### 2.2.2 Exclusion Criteria

1. Intracranial hemorrhage, tumor, trauma, or other neurological disorders on CT/MRI.
2. Posterior circulation or bilateral infarcts.
3. Prior intravenous thrombolysis or mechanical thrombectomy.
4. Contraindications to CT/MRI.
5. Incomplete imaging/clinical data or poor image quality.

### 2.3 Imaging Protocol

All patients underwent one-stop CTA-CTP scanning using the United Imaging uCT960+ system and MRI examination with diffusion-weighted imaging (DWI) sequences performed on a GE 1.5T HD-xt scanner equipped with an 8-channel head coil. Prior to imaging, an intravenous catheter was placed in the right median cubital vein, and patients received standardized instructions regarding examination procedures and potential physiological reactions during scanning.

All patients underwent one-stop CT scanning using the United Imaging uCT960+ 640-slice CT scanner (320 detector rows), which included sequential non-contrast CT, CT perfusion (CTP), and CT angiography (CTA) scans. The non-contrast CT parameters were set at 120 kV tube voltage, 300 mA tube current, with 5 mm slice thickness and 5 mm slice interval. For CTP scanning, the parameters were 100 kV tube voltage, 120 mA tube current, 5 mm slice thickness and interval, 1.0 s rotation time, using a power injector to administer 50 mL of non-ionic iodinated contrast (Iohexol, GE Healthcare, 350 mgI/mL) followed by 30 mL saline flush at 5 mL/s injection rate through the right median cubital vein, with a 5 s delay before initiating scanning over a 160 mm range from skull base to vertex, acquiring 19 dynamic scans. The acquired data were reconstructed into 5 mm thick slices with 5 mm intervals for CTP analysis and 1.5 mm slice width for multiphase CTA (mCTA) analysis, with mCTA images generated by post-processing the original CTP data using the United Imaging CT workstation. Multimodal MRI was performed on a GE 1.5T HD-xt system (GE Healthcare, USA) with the following protocol: T1-weighted imaging (TR/TE=1625/24 ms, FOV 240×240 mm, 6 mm slice thickness, acquisition time 1 min 14 s), T2-weighted imaging (TR/TE=4949/108.7 ms, FOV 240×240 mm, 6 mm slice thickness, 45 s acquisition), diffusion-weighted imaging (TR/TE=4053/84.2 ms, b-value=1000 s/mm^2^, FOV 240 × 240 mm, 6 mm slice thickness, 32 s acquisition), and FLAIR imaging (TR/TE=8400/100 ms, 5 mm slice thickness, FOV 240×240 mm, 2 min 23 s acquisition).

### 2.4 Image Analysis

All images were independently evaluated by two experienced neuroradiologists blinded to clinical data, with slice-by-slice analysis of lesions across all sequences. Discrepancies were resolved through consensus discussion.

CTP raw data were processed using uAI Discover – CerebralPerfusion V1.0 (United Imaging Intelligence, China), with the contralateral ICA/MCA origin manually marked as input artery and superior sagittal sinus as output vein to generate time-density curves (TDC). Manual correction was applied when TDC abnormalities resulted from incorrect vessel selection. The software automatically produced color-coded perfusion maps (CBF, CBV, MTT, TTP, Tmax) and a bone-removed CTA-MIP image from the optimal phase. CTA images were reconstructed into MPR, MIP, and VR formats. Multiphase CTA data were analyzed using FastStroke software, with two neuroradiologists assessing collateral circulation status on triphasic CTA-MIP and sCTA-MIP images, using the contralateral hemisphere as reference.

All imaging analyses were performed using standardized scoring systems. The Alberta Stroke Program Early CT Score (ASPECTS) was applied to DWI images, dividing the MCA territory into 10 regions across nuclear and supra-nuclear levels: M1 (anterior MCA cortex), M2 (lateral insula), M3 (posterior MCA cortex) at the nuclear level; M4-M6 (anterior, lateral and posterior MCA cortices) at the supra-nuclear level; plus caudate head (C), lentiform nucleus (L), internal capsule posterior limb (IC), and insula (I). Scores range from 10 (no infarction) to 0 (complete MCA territory infarction) [13].

Fluid-attenuated inversion recovery vascular hyperintensities (FVH) were defined as serpentine, tubular or dot-like hyperintensities along vessels on FLAIR sequences. Using modified ASPECTS regions (insula [I] plus M1-M6), each FVH-positive region scored 1 point (range: 0-7) [14].

The Maas collateral score evaluated sylvian fissure and cortical collateral filling: 1=absent; 2=reduced; 3=equal; 4=greater; 5=complete collateral flow. Scores 1-2 indicated poor and 3-5 good collaterals [15].

The regional leptomeningeal collateral (rLMC) score assessed collateral filling in ASPECTS regions (M1-M6), ACA territory, basal ganglia and sylvian fissure: 0=no filling; 1=reduced; 2=equal/greater (sylvian fissure scored 0/2/4). Total scores 0-10 indicated poor and 11-20 good collaterals [16].

The ASITN/SIR collateral grading system (modified from multiphase CTA-DSA) classified collateral flow as: 0=no collaterals; 1=slow peripheral filling with persistent defect; 2=rapid peripheral filling with partial ischemic zone perfusion; 3=slow but complete late venous filling; 4=complete rapid retrograde perfusion. Grades 0-2 indicated poor and 3-4 good collaterals [17].

**Figure 1.**
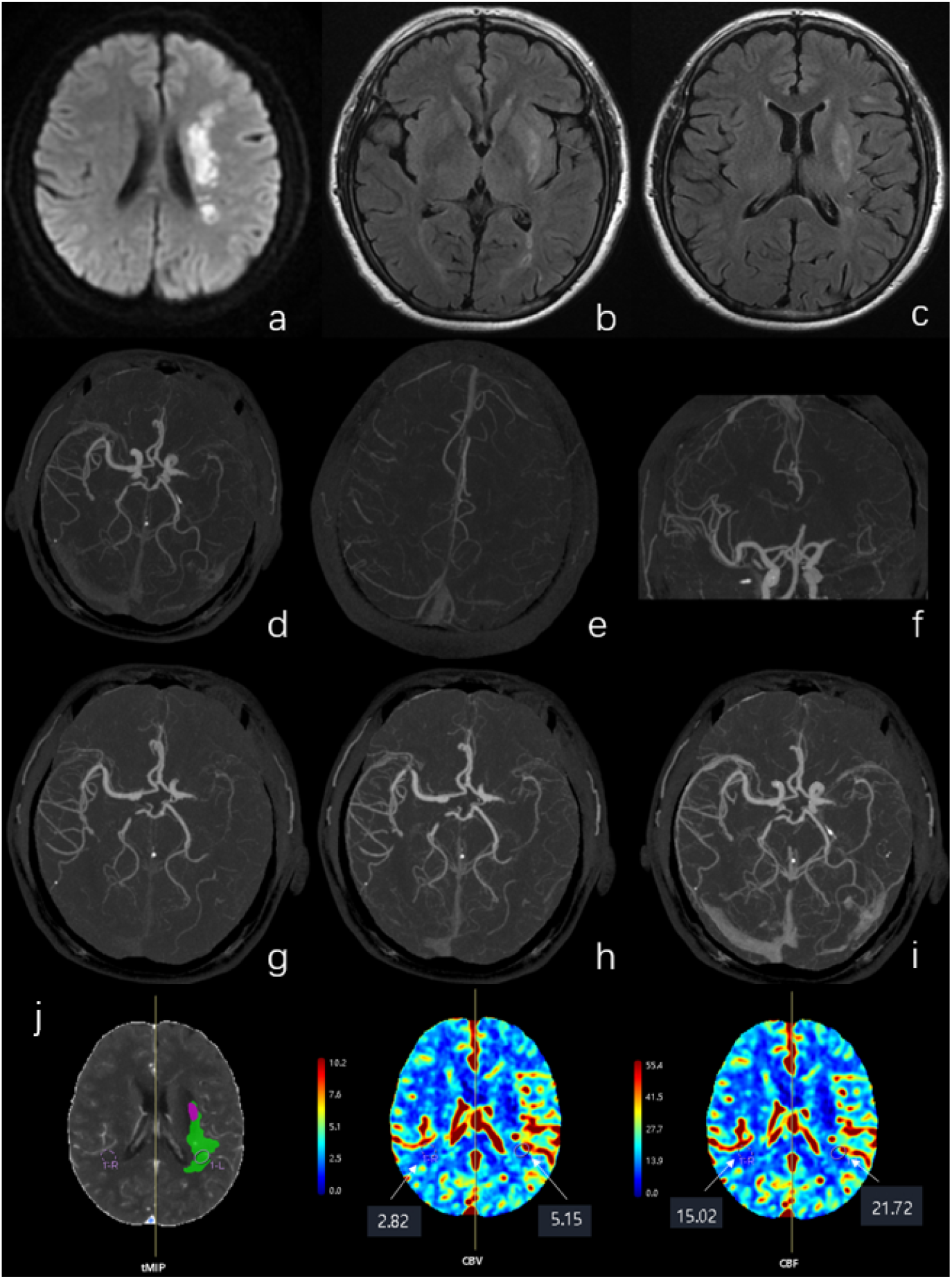
This is a patient with occlusion of the M1 segment of the MCA, with a 90-day mRS score of 2. (a) shows the patient’s DWI revealing acute cerebral infarction. (b) and (c) are T2 FLAIR images demonstrating the FVH sign. (d), (e), and (f) are SCTA images confirming occlusion of the left MCA. (g), (h), and (i) are reconstructed mCTA images, showing only partial collateral blood flow to the ischemic region, with persistent perfusion deficits. (j) indicates that the patient’s affected side exhibits higher CBF and CBV compared to the unaffected side. MCA: Middle Cerebral Artery; DWI: Diffusion-Weighted Imaging; FVH: FLAIR Vascular Hyperintensity; sCTA: Spiral CT Angiography; mCTA: Multiphase CT Angiography; CBV: Cerebral Blood Volume; CBF: Cerebral Blood Flow

### 2.5 Statistical Analysis

Statistical analyses were performed using R software (version 4.2.0). Continuous variables with normal distribution were expressed as mean ± standard deviation (SD), and group comparisons were conducted using Student’s *t*-test. For non-normally distributed continuous variables, data were presented as median (interquartile range, IQR), and the Wilcoxon rank-sum test was applied for group comparisons. Categorical variables were described as frequencies (percentages), and differences between groups were assessed using the chi-square test or Fisher’s exact test, as appropriate.

Inter-rater reliability for CT- and MRI-based collateral circulation scoring scales was evaluated using Cohen’s kappa (κ) statistics. Spearman’s rank correlation analysis was performed to assess the association between CTA/MRI-derived collateral scores and the ASITN/SIR collateral grading system (modified from multiphase CTA-DSA).

Univariate analysis followed by multivariate binary logistic regression was used to identify independent predictors of patient prognosis (favorable vs. poor outcome). Receiver operating characteristic (ROC) curve analysis was employed to evaluate the predictive performance of collateral circulation scores for clinical outcomes in conservatively treated patients. A two-tailed *P*-value < 0.05 was considered statistically significant.

## 3. Results

### 3.1. Clinical Outcomes

A total of 112 patients were ultimately included in the final analysis after applying the predefined inclusion/exclusion criteria. Exclusions comprised 18 cases with onset-to-examination intervals exceeding 72 hours, 6 cases demonstrating concurrent intracranial hemorrhage, 27 cases receiving thrombolytic or thrombectomy treatment prior to imaging, 51 cases presenting with posterior circulation or bilateral infarctions, 1 case excluded due to renal insufficiency precluding CTA-CTP completion, and 9 cases eliminated because of missing imaging data or inadequate image quality for proper evaluation.

**Figure 2.**
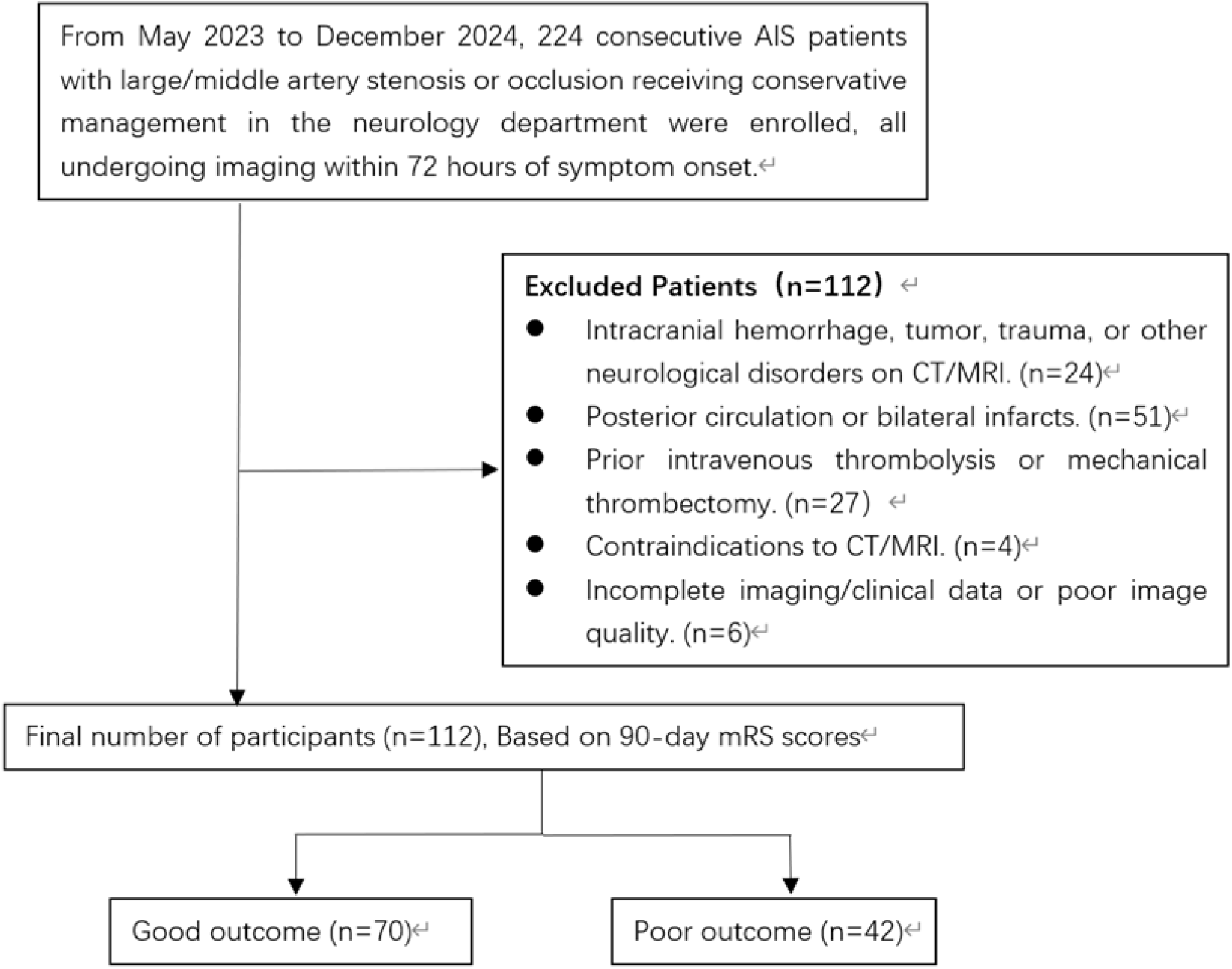
Flow diagram of enrollment of study patients

**Table 1.**
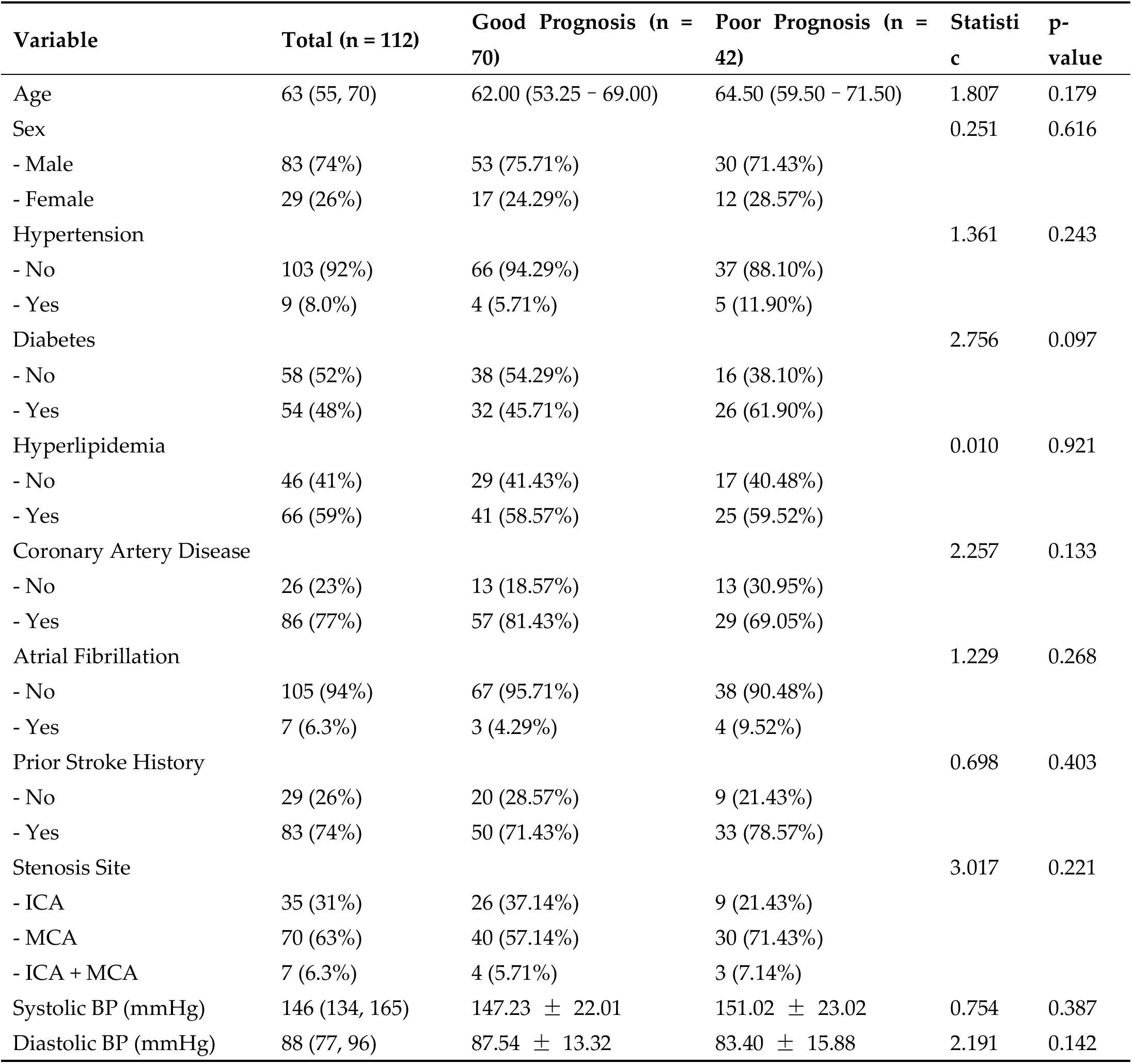
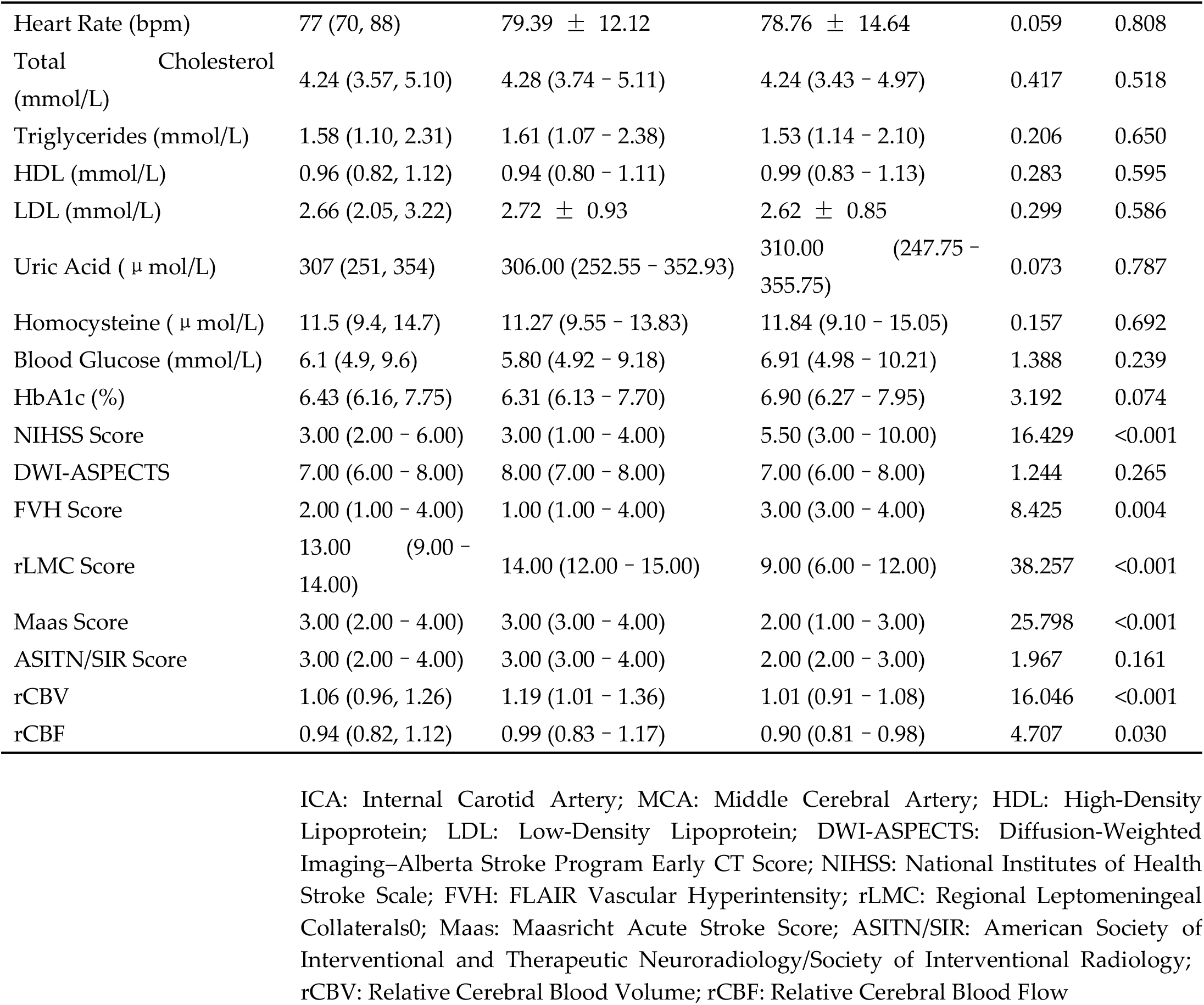
Comparisons of clinical and imaging characteristics in patients with good and poor outcome.

The study cohort comprised 112 AIS patients with a median age of 63 years (interquartile range [IQR] 55-70), including 83 males (74%) and 29 females (26%). Comorbid conditions were present as follows: hypertension in 103 patients (92%), diabetes mellitus in 58 (52%), hyperlipidemia in 46 (41%), coronary artery disease in 26 (23%), atrial fibrillation in 7 (6.3%), and previous stroke history in 29 (26%). Admission vital signs demonstrated a median systolic blood pressure of 146 mmHg (IQR 134-165), diastolic pressure of 88 mmHg (IQR 77-96), and heart rate of 77 beats per minute (IQR 70-88). Laboratory parameters at admission included a median blood glucose level of 6.1 mmol/L (IQR 4.9-9.6), glycated hemoglobin of 6.43% (IQR 6.16-7.75), homocysteine of 11.5 μmol/L (IQR 9.4-14.7), total cholesterol of 4.24 mmol/L (IQR 3.57-5.10), triglycerides of 1.58 mmol/L (IQR 1.10-2.31), high-density lipoprotein of 0.96 mmol/L (IQR 0.82-1.12), low-density lipoprotein of 2.66 mmol/L (IQR 2.05-3.22), and uric acid of 307 μmol/L (IQR 251-354). Vascular imaging revealed internal carotid artery stenosis in 35 patients (31.3%), middle cerebral artery stenosis in 70 (62.5%), and combined lesions in 7 (6.3%). Neurological assessment showed a median NIHSS score of 3 (IQR 2-6) and DWI-ASPECTS of 7 (IQR 6-8) at admission.

### 3.2 Collateral Circulation Scoring Results in AIS Patients

The evaluation of collateral circulation using various scoring systems demonstrated the following distribution among the 112 AIS patients:

For CTA-based collateral assessments, the Maas score classified 46 patients (41.1%) as having poor collateral circulation (scores 1-2) and 66 patients (58.9%) with good collateral circulation (scores 3-5). The rLMC scoring system identified 43 patients (38.4%) with poor collaterals (scores 0-10) and 69 patients (61.6%) with good collaterals (scores 11-20). When applying the ASITN/SIR grading system adapted from DSA standards, 40 patients (35.7%) showed poor collateral circulation (grades 0-2) while 72 patients (64.3%) exhibited good collateral circulation (grades 3-4).

The FVH scoring system based on FLAIR MRI sequences categorized patients into 57 cases (50.9%) with lower scores (0-3 points) and 49 cases (43.8%) with higher scores (4-7 points).

### 3.3 Inter-rater Reliability of Collateral Circulation Scores

The Kappa consistency analysis revealed good agreement between junior and senior radiologists across all three collateral scoring systems. The inter-rater reliability was highest for the rLMC score, followed by FVH score, with the Maas score showing slightly lower but still acceptable consistency between raters.

**Table 2.**
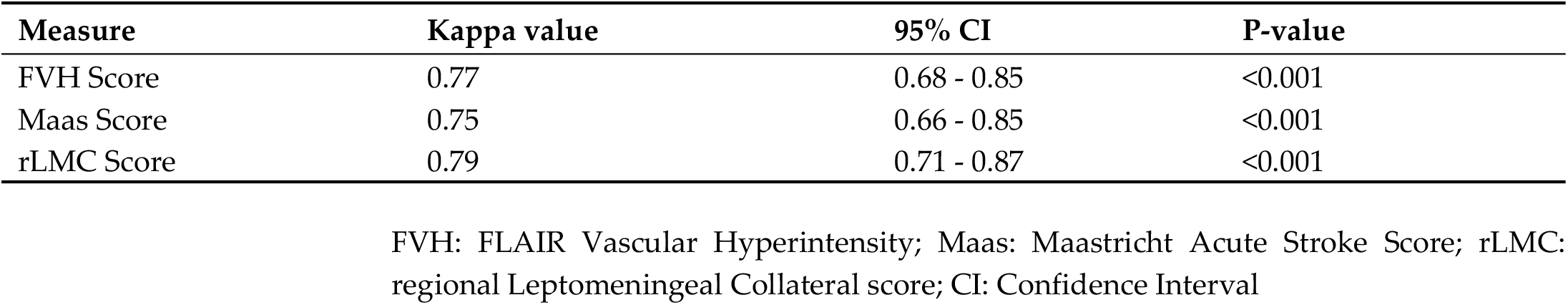
Inter-rater reliability of collateral circulation scoring.

| Measure | Kappa value | 95% CI | P-value |
| --- | --- | --- | --- |
| FVH Score | 0.77 | 0.68 - 0.85 | <0.001 |
| Maas Score | 0.75 | 0.66 - 0.85 | <0.001 |
| rLMC Score | 0.79 | 0.71 - 0.87 | <0.001 |

FVH: FLAIR Vascular Hyperintensity; Maas: Maastricht Acute Stroke Score; rLMC: regional Leptomenigeal Collateral score; CI: Confidence Interval

### 3.4 Correlation Analysis Between Collateral Scores and ASITN/SIR Grading System

Spearman correlation analysis demonstrated that rLMC score and Maas score were positively correlated with ASITN/SIR collateral grading, while FVH score showed a negative correlation (p < 0.001). The correlation coefficients, ranked from highest to lowest, were as follows: rLMC score > Maas score > FVH score.

**Table 3.**
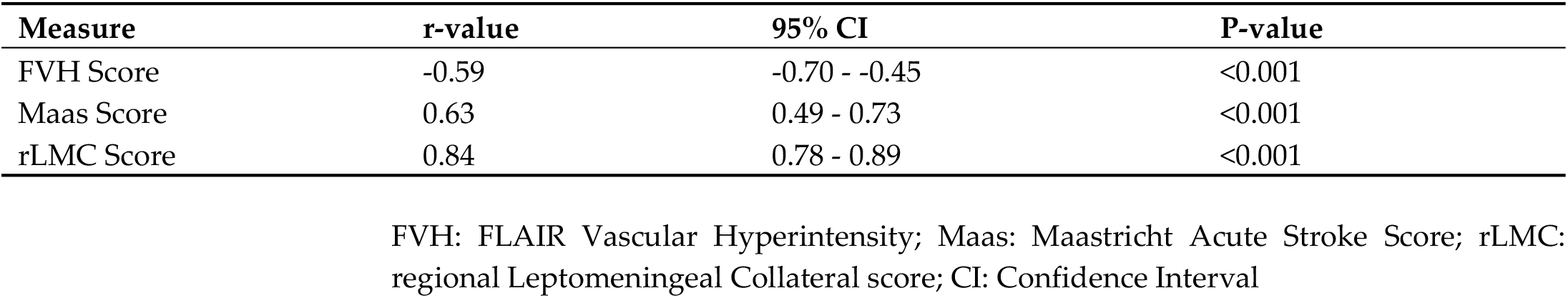
Correlation Analysis Between Collateral Scores and ASITN/SIR Grading System.

### 3.5 Analysis of Prognostic Factors in AIS Patients

There were 70 patients with favorable outcomes and 42 patients with poor outcomes. Univariate analysis of the overall sample showed that compared with patients having poor outcomes, those with favorable outcomes had significantly lower admission NIHSS scores (p<0.05), significantly lower FVH scores (p<0.05), significantly higher proportions of good collateral circulation status as indicated by rLMC and Maas scores (p<0.05), and significantly higher rCBV and rCBF values (p<0.05). No statistically significant differences were found between the two groups for all other indicators (all p>0.05).

Variables with p<0.05 in univariate analysis were included as independent variables in the multivariate logistic regression model. The results demonstrated that admission NIHSS score (OR 5.09, 95% CI 1.44-17.96, p=0.011), FVH score (OR 4.31, 95% CI 1.26-14.67, p=0.020), rLMC score (OR 0.11, 95% CI 0.02-0.52, p=0.005), and rCBV (OR 5.40, 95% CI 1.03-28.35, p=0.046) were independent influencing factors for AIS patient prognosis (all p<0.05). Although rCBF was found to be associated with poor outcomes in univariate analysis, no significant association was detected between rCBF (OR 0.81, 95% CI 0.17-3.86, p=0.788) and outcomes in multivariate analysis.

**Table 4.** Evaluation of factors associated with prognosis by univariate logistic regression analysis.

| Variable | Good Outcome (N=70) | Poor Outcome (N=42) | OR (95% CI) | p-value |
| --- | --- | --- | --- | --- |
| Sex |  |  | 1.25 (0.53–2.96) | 0.617 |
| - Male | 53 (75.7%) | 30 (71.4%) |  |  |
| - Female | 17 (24.3%) | 12 (28.6%) |  |  |
| Age (years) | 61.2 ± 11.8 | 64.1 ± 9.6 | 1.03 (0.99–1.06) | 0.179 |
| NIHSS Score |  |  |  |  |
| - ≤3 | 48 (68.6%) | 11 (26.2%) |  |  |
| - >3 | 22 (31.4%) | 31 (73.8%) | 6.15 (2.62–14.43) | <0.001 |
| Hypertension |  |  |  |  |
| - No | 4 (5.7%) | 5 (11.9%) |  |  |
| - Yes | 66 (94.3%) | 37 (88.1%) | 0.45 (0.11–1.77) | 0.253 |
| Diabetes |  |  |  |  |
| - No | 38 (54.3%) | 16 (38.1%) |  |  |
| - Yes | 32 (45.7%) | 26 (61.9%) | 1.93 (0.88–4.21) | 0.099 |
| Hyperlipidemia |  |  |  |  |
| - No | 41 (58.6%) | 25 (59.5%) |  |  |
| - Yes | 29 (41.4%) | 17 (40.5%) | 0.96 (0.44–2.09) | 0.921 |
| Coronary Artery Disease |  |  |  |  |
| - No | 57 (81.4%) | 29 (69.0%) |  |  |
| - Yes | 13 (18.6%) | 13 (31.0%) | 1.97 (0.81–4.78) | 0.136 |
| Atrial Fibrillation |  |  |  |  |
| - No | 67 (95.7%) | 38 (90.5%) |  |  |
| - Yes | 3 (4.3%) | 4 (9.5%) | 2.35 (0.50–11.06) | 0.279 |
| Prior Stroke |  |  |  |  |
| - No | 50 (71.4%) | 33 (78.6%) |  |  |
| - Yes | 20 (28.6%) | 9 (21.4%) | 0.68 (0.28–1.68) | 0.405 |
| Stenosis Location |  |  | 2.17 (0.40–11.60) | 0.366 |
| - ICA | 26 (37.1%) | 9 (21.4%) |  |  |
| - MCA | 40 (57.1%) | 30 (71.4%) |  |  |
| - ICA+MCA | 4 (5.7%) | 3 (7.1%) |  |  |
| DWI-ASPECTS |  |  |  |  |
| - ≤7 | 34 (48.6%) | 26 (61.9%) |  |  |
| - >7 | 36 (51.4%) | 16 (38.1%) | 0.58 (0.27–1.27) | 0.172 |
| FVH Score |  |  |  |  |
| - ≤2.5 | 49 (70.0%) | 8 (19.0%) |  |  |
| - >2.5 | 21 (30.0%) | 34 (81.0%) | 9.92 (3.93–24.99) | <0.001 |
| rLMC Score |  |  |  |  |
| - ≤11 | 13 (18.6%) | 30 (71.4%) |  |  |
| - >11 | 57 (81.4%) | 12 (28.6%) | 0.09 (0.04–0.22) | <0.001 |
| Maas Score |  |  |  |  |
| - ≤2 | 17 (24.3%) | 29 (69.0%) |  |  |
| - >2 | 53 (75.7%) | 13 (31.0%) | 0.14 (0.06–0.34) | <0.001 |
| ASITN Score | 2.9 ± 0.8 | 3.1 ± 1.1 | 1.31 (0.86–2.00) | 0.214 |
| rCBV |  |  |  |  |
| - >1.11 | 40 (57.1%) | 8 (19.0%) |  |  |
| - ≤1.11 | 30 (42.9%) | 34 (81.0%) | 5.67 (2.29–13.99) | <0.001 |
| rCBF |  |  |  |  |
| - ≤0.97 | 31 (44.3%) | 32 (76.2%) |  |  |
| - >0.97 | 39 (55.7%) | 10 (23.8%) | 0.25 (0.11–0.58) | 0.001 |
| Systolic BP (mmHg) | 147.2 ± 22.0 | 151.0 ± 23.0 | 1.01 (0.99–1.03) | 0.384 |
| Diastolic BP (mmHg) | 87.5 ± 13.3 | 83.4 ± 15.9 | 0.98 (0.95–1.01) | 0.145 |
| Heart Rate (bpm) | 79.4 ± 12.1 | 78.8 ± 14.6 | 1.00 (0.97–1.03) | 0.806 |
| Total Cholesterol (mmol/L) | 8.8 ± 36.8 | 4.3 ± 1.0 | 0.90 (0.62–1.29) | 0.554 |
| Triglycerides (mmol/L) | 1.9 ± 1.3 | 1.8 ± 1.0 | 0.88 (0.62–1.24) | 0.45 |
| HDL (mmol/L) | 1.0 ± 0.3 | 1.0 ± 0.2 | 1.28 (0.26–6.37) | 0.762 |
| LDL (mmol/L) | 2.7 ± 0.9 | 2.6 ± 0.8 | 0.88 (0.57–1.35) | 0.554 |
| Uric Acid (μmol/L) | 318.6 ± 88.1 | 304.1 ± 76.8 | 1.00 (0.99–1.00) | 0.377 |
| Homocysteine ($\mu\text{mol/L}$) | $13.3 \pm 7.6$ | $13.4 \pm 7.2$ | 1.00 (0.95-1.06) | 0.912 |
| Blood Glucose ($\text{mmol/L}$) | $7.1 \pm 2.9$ | $8.0 \pm 3.6$ | 1.09 (0.97-1.23) | 0.137 |
| HbA1c (%) | $6.9 \pm 1.6$ | $7.2 \pm 1.2$ | 1.18 (0.90-1.55) | 0.239 |

NIHSS: National Institutes of Health Stroke Scale; ICA: Internal Carotid Artery; MCA: Middle Cerebral Artery; DWI-ASPECTS: Diffusion-Weighted Imaging–Alberta Stroke Program Early CT Score; FVH: FLAIR Vascular Hyperintensity; rLMC: Regional Leptomeningeal Collaterals; Maas: Maasricht Acute Stroke Score; ASITN/SIR: American Society of Interventional and Therapeutic Neuroradiology/Society of Interventional Radiology; rCBV: Relative Cerebral Blood Volume; rCBF: Relative Cerebral Blood Flow; HDL: High-Density Lipoprotein; LDL: Low-Density Lipoprotein;

**Table 5.** Multivariate logistic regression analysis.

|  | Estimate | SE | Wald | p | OR(95%CI) |
| --- | --- | --- | --- | --- | --- |
| (Intercept) | -1.863 | 0.952 | -1.958 | 0.05 | 0.155(0.024 - 1.002) |
| NIHSS Score >3 | 1.628 | 0.643 | 2.532 | 0.011 | 5.093(1.445 - 17.955) |
| FVH Score >2.5 | 1.46 | 0.625 | 2.334 | 0.02 | 4.305(1.264 - 14.670) |
| rLMC Score >11 | -2.244 | 0.808 | -2.779 | 0.005 | 0.106(0.022 - 0.516) |
| Maas Score >2 | -0.309 | 0.796 | -0.388 | 0.698 | 0.734(0.154 - 3.498) |
| rCBV $\leq 1.11$ | 1.686 | 0.846 | 1.991 | 0.046 | 5.396(1.027 - 28.351) |
| rCBF >0.97 | -0.215 | 0.799 | -0.269 | 0.788 | 0.806(0.168 - 3.864) |

SE: Standard Error; OR: Odds Ratio; CI: Confidence Interval; NIHSS: National Institutes of Health Stroke Scale; FVH: FLAIR Vascular Hyperintensity; rLMC: Regional Leptomeningeal Collaterals; Maas: Maasricht Acute Stroke Score; ASITN/SIR: American Society of Interventional and Therapeutic Neuroradiology/Society of Interventional Radiology; rCBV: Relative Cerebral Blood Volume; rCBF: Relative Cerebral Blood Flow;

### 3.6 ROC Curve Analysis of Factors Associated with Poor Prognosis

ROC curve analysis demonstrated that the AUC values of each indicator were ranked from high to low as follows: rLMC score, rCBV, admission NIHSS score, and FVH score (all p<0.05). The optimal cutoff values were: 11.5 for rLMC score (sensitivity 0.810, specificity 0.700); 3.5 for NIHSS score (sensitivity 0.687, specificity 0.686); 1.111 for rCBV (sensitivity 0.571, specificity 0.810); and 2.5 for FVH score (sensitivity 0.810, specificity 0.700). Among these indicators, the Youden index corresponding to the optimal cutoff value of rLMC score (0.528) was higher than that of other variables. The predictive value of individual indicators for poor prognosis was ranked as: rLMC score > rCBV > admission NIHSS score > FVH score.

**Figure 3.**
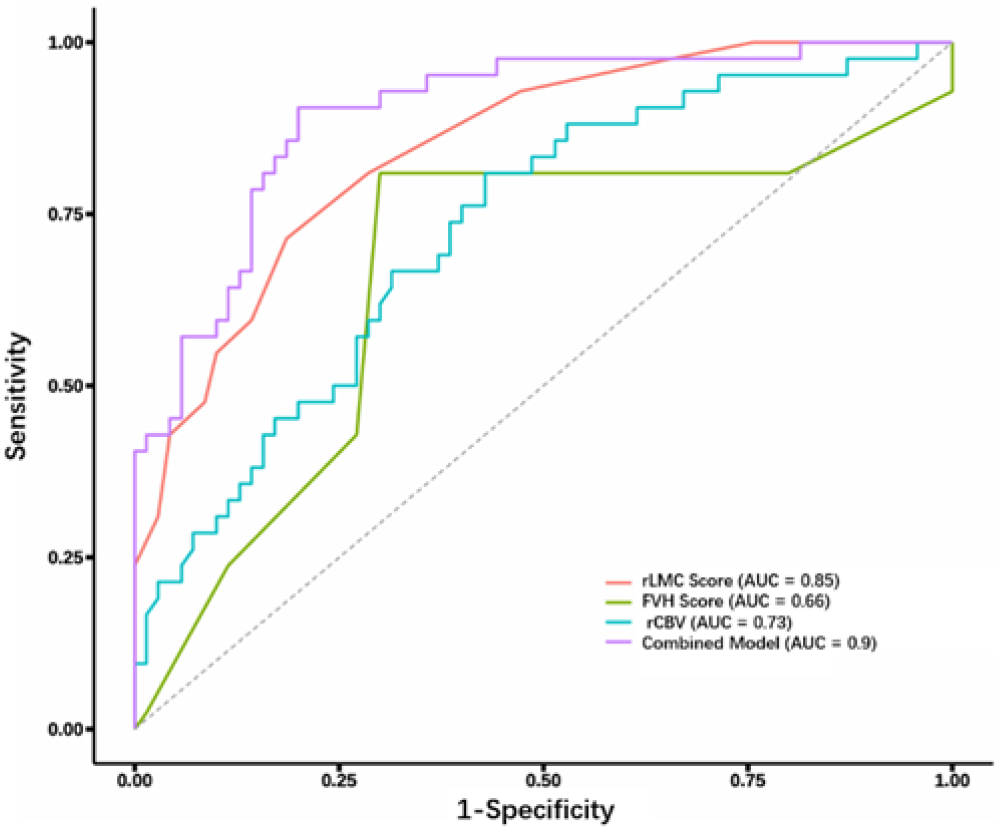
ROC analysis of FVH, rLMC, and rCBV individually and in combination for predicting poor outcomes ROC: Receiver operating characteristic; FVH: FLAIR Vascular Hyperintensity; rLMC: Regional Leptomeningeal Collaterals; rCBV: Relative Cerebral Blood Volume; AUC: area under the curve;

**Table 6.** The area under the ROC curve (AUC) of various parameters for predicting poor prognosis in AIS.

| Variable | AUC (95% CI) | Sensitivity (95% CI) | Specificity (95% CI) | Cut-off | Youden's Index |
| --- | --- | --- | --- | --- | --- |
| NIHSS Score | 0.727 (0.628–0.826) | 0.667 (0.524–0.809) | 0.686 (0.577–0.794) | >3.5 | 0.353 |
| FVH Score | 0.662 (0.550–0.774) | 0.810 (0.691–0.928) | 0.700 (0.593–0.807) | >2.5 | 0.51 |
| rLMC Score | 0.848 (0.778–0.919) | 0.814 (0.723–0.905) | 0.714 (0.578–0.851) | ≤11.5 | 0.528 |
| rCBV | 0.727 (0.631–0.822) | 0.571 (0.455–0.687) | 0.810 (0.691–0.928) | ≤1.111 | 0.381 |

NIHSS: National Institutes of Health Stroke Scale; FVH: FLAIR Vascular Hyperintensity; rLMC: Regional Leptomeningeal Collaterals; rCBV: Relative Cerebral Blood Volume; AUC: area under the curve; CI: Confidence Interval;

## 4. Discussion

Intravenous thrombolysis remains the most effective pharmacological treatment for improving outcomes in acute ischemic stroke. However, the clinical application of the most commonly used thrombolytic agent, alteplase, is limited by its narrow therapeutic window of 4.5 hours [18], while urokinase, now less frequently used, has a slightly extended but still restrictive 6-hour window [19]. Various factors often prevent patients from receiving timely thrombolytic therapy within these critical timeframes, significantly constraining its clinical utility. Recent clinical studies have demonstrated that robust collateral circulation can effectively limit the expansion of the infarct core, enhance clinical outcomes, and reduce the risk of disease recurrence [20]. Consequently, the establishment of collateral circulation, particularly through leptomeningeal arteries, has emerged as a crucial independent factor influencing the prognosis of ischemic stroke patients [21].

Collateral circulation refers to the anastomotic vascular structures connecting adjacent arterial branches, present in most human tissues. Under physiological conditions, these structures regulate blood flow to meet tissue metabolic demands. In pathological scenarios such as severe stenosis or occlusion of major arteries, these anastomotic channels can establish alternative blood flow pathways to maintain perfusion to ischemic tissues [22].

In previous research, the ASITN/SIR scoring system has been established as the gold standard for cerebrovascular assessment due to its internationally validated reliability through multicenter clinical trials [17,23]. This system combines the high spatial resolution of digital subtraction angiography (capable of visualizing even the smallest leptomeningeal vessels) with real-time dynamic imaging, providing clinicians with comprehensive multidimensional information about blood flow patterns, velocity, and vascular morphology [24]. Our study evaluated the correlations between rLMC score, Maas score, FVH score, and ASITN/SIR grading. The experimental data demonstrated that rLMC score showed the highest correlation coefficient with ASITN/SIR grading (p<0.001) compared to other scoring systems.

Numerous studies have indicated that FVH formation is closely associated with leptomeningeal collateral compensation [9,11,25,26]. The rLMC score quantitatively assesses collateral circulation function by evaluating the extent of collateral vessel recruitment and compensatory range. Our findings support the view that FVH score can serve as a quantitative indicator of collateral compensation capacity. This correlation may exist because FVH scoring, by counting hyperintense vessels on FLAIR sequences, indirectly reflects the total number of recruited collateral vessels during acute ischemia. Furthermore, the hyperintensity resulting from absent flow voids due to sluggish blood flow may correlate with the severity of hemodynamic impairment. Similarly, CTA-based collateral grading evaluates visible collateral vessels, providing numerical assessment of collateral function from a different imaging modality. When leptomeningeal collaterals are partially recruited (low rLMC score) but fail to maintain effective perfusion due to increased microvascular resistance or blood-brain barrier disruption, FVH may indicate unsuccessful compensation attempts rather than true reperfusion.

In this retrospective study, we observed a moderate negative correlation between FVH and CTA-based collateral scores, suggesting some association between these parameters. However, this correlation did not reach statistical significance, possibly due to limited sample size and potential confounding factors such as reperfusion time and infarct core progression rate. Indeed, beyond collateral grading, FVH may be influenced by other factors that should be considered in future neuroimaging models of stroke outcomes.

Our study systematically analyzed key factors affecting outcomes in conservatively treated acute ischemic stroke (AIS) patients by integrating multimodal imaging parameters and clinical indicators. The results revealed that patients with poor outcomes typically exhibited: 1) significantly higher admission NIHSS scores; 2) more extensive FVH hyperintensity on FLAIR sequences; 3) markedly reduced rLMC collateral scores on CTA-CTP; and 4) significantly decreased cerebral blood volume (CBV) values in affected regions.

The admission NIHSS score emerged as an independent predictor of AIS patient outcomes, with higher scores associated with poorer prognosis. Widely used in clinical practice, NIHSS provides preliminary assessment of neurological function, which is closely related to perfusion compensation in ischemic regions - a key determinant of patient outcomes. This finding aligns with previous studies identifying admission NIHSS as an independent prognostic factor for AIS patients [22,27].

The rLMC score, a quantitative CTA-based evaluation method, offers comprehensive coverage of six ASPECTS regions in the middle cerebral artery territory, anterior cerebral artery region, and basal ganglia, with particular emphasis on sylvian fissure artery scoring (0/2/4 points) [16,28]. Compared to other collateral scoring systems (such as Tan score or Maas score), rLMC provides more detailed regional assessment, better reflecting collateral compensation after large vessel occlusion. The degree of collateral compensation in these regions significantly correlates with reduced infarct volume [29]. Our multimodal imaging analysis confirmed that regional leptomeningeal collateral (rLMC) score is an independent factor predicting 90-day functional outcomes in AIS patients. Multivariate logistic regression showed that patients with rLMC scores >11 had significantly lower risk of poor outcomes (OR=0.11, 95%CI 0.02-0.52, p=0.005), consistent with Menon et al.’s findings [16], while extending clinical applications through combination with FVH and CTP parameters.

The mechanisms underlying FVH formation remain complex and controversial. Current evidence suggests FVH is closely related to hemodynamic changes following acute large vessel occlusion, but its clinical interpretation requires consideration of multiple factors including occlusion severity, collateral status, and therapeutic interventions. The prognostic reliability of FVH varies significantly across studies due to heterogeneous mechanisms and complex relationships with ischemic injury [30]. Previous research presents several perspectives: 1) FVH may correlate with favorable outcomes, as some studies show positive association between FVH scores and collateral status [26,31]; 2) High FVH scores may indicate more severe hypoperfusion and neurological deficits at admission, potentially predicting poor outcomes [14,25,32,33]; and 3) FVH may have no prognostic value, as Jing et al. [34] concluded that despite representing slow or reversed collateral flow, FVH fails to provide compensatory benefit. While opinions vary, consensus exists that extensive FVH is associated with severe stenosis or occlusion of large vessels [31]. Our multivariate logistic regression analysis demonstrated that higher FVH scores and lower collateral scores independently correlated with poor outcomes. ROC analysis further confirmed FVH’s excellent predictive performance for conservative treatment outcomes in AIS patients. Additionally, our correlation analysis showed baseline FVH scores could reflect stroke severity across multiple post-treatment stages, supporting FVH-ASPECT scoring as a stable predictor of conservative treatment efficacy. In summary, pretreatment FVH assessment provides valuable and essential information for predicting functional outcomes and survival rates.

In our study, rCBV emerged as another independent predictor of 90-day poor outcomes. Results showed significantly higher rCBV values in the favorable outcome group (mRS 0-2) compared to the poor outcome group (mRS 3-6) (p<0.05), confirmed by multivariate logistic regression. This finding aligns with previous research [35,36], as CBV closely relates to collateral status and reflects compensatory capacity for hemodynamic changes. CBV serves as a crucial parameter for assessing whether cerebral autoregulation remains functional after ischemia [37,38]. Reduced CBV typically indicates microcirculatory impairment in local brain tissue. In AIS caused by hypoperfusion-induced cerebral hypoxia, higher CBV values suggest adequate tissue oxygenation and lower risk of microcirculatory dysfunction affecting prognosis [39]. Significant CBV reduction effectively identifies and evaluates stroke severity, particularly in distinguishing normal from ischemic brain tissue [40]. Our findings confirm the association between reduced rCBV and poor outcomes, while demonstrating the synergistic value of combining FVH scoring with multimodal CT parameters (rLMC score, rCBV) for predicting collateral status and functional outcomes. The combined model’s predictive performance (AUC=0.9) significantly surpassed individual indicators, overcoming limitations of single-modality assessment. These findings provide valuable clinical references for patient management and outcome prediction.

This study has several limitations. As a single-center retrospective analysis including only conservatively treated patients within 72 hours of onset, while excluding those receiving intravenous thrombolysis or mechanical thrombectomy, potential selection bias exists. The relatively small sample size also warrants caution. Future multicenter prospective studies with larger cohorts are needed to further validate the correlation between FVH and CTA-based collateral scores and their predictive value for conservative treatment outcomes.

## 5. Conclusions

In conclusion, admission NIHSS score, FVH score, rLMC score, and rCBV may serve as independent predictors for conservative treatment outcomes in AIS patients, with FVH score negatively correlating with collateral scores. Specifically, high FVH scores combined with low rLMC scores and low rCBV values correlate with poor outcomes, and their combined use enhances prediction accuracy. The integration of multimodal CT and MRI facilitates early AIS diagnosis and comprehensive collateral circulation assessment.

## Data Availability

The data that support the findings of this study are available from the corresponding author upon reasonable request.

## Author Contributions

Conceptualization: Q.Y. and X.Q.; methodology: Y.M., Q.Y. and Y.C.; software: X.Q. and H.S.; data curation: Q.Y. and Q.X.; investigation: Q.Y., Y.C. and H.S.; validation: G.Y. and Y.M.; formal analysis: Q.Y., and X.Q.; visualization: G.Y.; writing - original draft: Q.Y. and X.Q.; writing - review & editing: Q.Y., Y.M., and Y.C., project administration: Y.M.; supervision: Y.M. and G.Y. All authors have read and agreed to the published version of the manuscript.

## Institutional Review Board Statement

This retrospective study was approved by the Ethics Committee of Daqing Oilfield General Hospital (Approval No.: ZYAF/SC-07/02.0; Date: January 10, 2025). The requirement for informed consent was waived by the ethics committee due to the retrospective nature of the study, which involved anonymized analysis of existing imaging and clinical data without additional patient intervention or risk.

## Informed Consent Statement

Informed consent was waived for this study by the Ethics Committee of Daqing Oilfield General Hospital because the research utilized de-identified retrospective data, and obtaining individual consent was impracticable without compromising the scientific validity of the study.

## Data Availability Statement

Dataset may be available upon reasonable written request.

